# Psychological distress and work and social adjustment in the COVID-19 pandemic: a cross-Country analysis

**DOI:** 10.1101/2022.01.30.22270127

**Authors:** Walter Distaso, Ana V. Nikčević, Marcantonio M. Spada

## Abstract

We investigate the determinants of several measures of psychological distress and work and social adjustment, using data from a large survey covering six countries and three continents over the COVID-19 pandemic. Our analysis reveals substantial cross-country heterogeneity and identifies a strong effect of COVID-19 specific measures of distress onto generic ones, but not the other way around. The results confirm the importance of controlling for individual characteristics, which help explain some of the cross-country differences. Finally, they also highlight specific categories of individuals who have recorded extremely high levels of psychological distress.

## 1 Introduction

On the 11^th^ of March 2020, the Coronavirus disease (COVID-19) was declared a pandemic by the World Health Organisation (WHO). Within a year (so by March 2021) there were 115 million confirmed cases and over 2.5 million deaths worldwide attributable to COVID-19. From the beginning of the COVID-19 outbreak, reports and studies indicating the presence of elevated psychological distress started emerging (e.g., Brailovskaia et al., 2021; Cao et al., 2020; Chen et al., 2021; Duong, 2021; Kontoangelos et al., 2020; Lee, 2020; Mansueto et al., 2021; Shevlin et al., 2020; Wang et al., 2020; Wu, Styra, & Gold, 2020). The psychological distress experienced was extensive, including depression and anxiety (Huang & Zhao, 2020; Oh et al., 2021), obsessive-compulsive (Abba-Aji et al., 2020; Wheaton et al., 2021; Seçer & Ulaş, 2020) and post-traumatic stress symptoms (Johnson et al., 2020; Liu et al., 2020; Akbari et al., 2021) as well as health anxiety (Akbari et al., 2021; Özdin & Bayrak Özdin, 2020). Furthermore, increases in substance use (Czeisler et al., 2020; Akbari et al., 2020), suicidal ideation (Czeisler et al., 2020), loneliness and fatigue (Mansueto et al., 2021; Hou et al., 2020), loss and grief (Wallace et al., 2020; Zhai & Du, 2020), sleep problems (Duong, 2021; Jahrami et al., 2021), and reduced life satisfaction (Duong, 2021) were also observed. These broad findings align themselves to those reported during the SARS outbreak (Cheng et al., 2004) and in the 2009 H1N1 pandemic (Rubin et al., 2009; Wheaton et al., 2012).

In this paper we focus on the relationship between generic measures of psychological distress and COVID-19 specific ones, in order to fully capture the effect of the pandemic on the psychological wellbeing of individuals. The core question of interest is to check whether (and to what extent) generic measures of psychological distress have been influenced by specific measures related to COVID-19. In this respect, our analysis is not merely descriptive, but tries to establish and quantify a relationship between these variables. To accomplish this task, we have conducted an extensive survey across six different countries, namely China, Germany, Italy, Sweden, the United Kingdom, and the United States. This choice has not been random, but intended to cover countries and areas of the world that differed markedly with respect to the status of the diffusion of the pandemic and/or the strength and enforcement of the respective Governments’ policies to contain it.

Besides measures of psychological distress, the survey has also collected data on personality traits, life satisfaction, individual attitude to risk and different social, economic and demographic factors. Overall, we collected more than 100 variables for each participant. Therefore, the rich information contained in our dataset is best geared to highlight the direct conditional relationship between generic and COVID-19 specific measures of psychological distress, stripping out the effect of potentially confounding factors related to either the state of the diffusion of the pandemic in a given Country or to individual characteristics.

In this respect, we add novel results to the existing literature because our analysis: (1) extends over three continents and reveals substantial cross-country evidence; (2) is based on a large and representative sample of individuals; (3) crucially controls for country specific effects and several individual social, economic and demographic factors; (4) and addresses the issue of potential endogeneity between the different measures of psychological distress.

Our results identify a clear channel of transmission from COVID-19 specific to generic measures of psychological distress and highlight categories of individuals that have been mostly hit in terms of a stronger conditional effect.

Given the social and economic implications of increased levels of psychological distress, we believe our results provide helpful evidence to devise and shape appropriate and effective government policies.

The paper is organized as follows. In Section 2 we describe the construction and main features of our extensive dataset. In Section 3 we present the results of our empirical analysis and in Section 4 we draw conclusions on our findings. All the Tables are reported in Appendix A.

## 2 Dataset construction

### 2.1 Participants’ characteristics

A total sample of 5,179 participants took part in the study and completed a battery of questionnaires, during March 2021. Participants were required to: (1) be at least 18 years of age; and (2) consent to participate. For each country where data was collected, study questionnaires were administered in the native language and a representative sample aged 18+ stratified by age, gender, and ethnicity was constructed.^1^

Table 1 presents key participants’ characteristics.^2^ We now describe the main variables contained in the survey.

**Table 1:** Dataset descriptive statistics.

|  | China | Germany | Italy | Sweden | UK | USA |
| --- | --- | --- | --- | --- | --- | --- |
| Participant numbers | 1560 | 510 | 529 | 503 | 541 | 1536 |
| Age (average) | 34.5 | 41.35 | 42.78 | 40.42 | 40.44 | 40.02 |
| Gender split (females) | 0.5 | 0.5 | 0.49 | 0.49 | 0.48 | 0.5 |

### 2.2 Measures

#### 2.2.1 Sociodemographic variables

Participants were asked to state their age, gender, ethnicity, marital status, number of children, number of people in household, employment status, employment type, employment category, education level, COVID-19 risk status, COVID-19 vaccination status, whether someone they knew has passed away due to COVID-19, weight, household income and wealth, and existing diagnosis of mental health disorder.

#### 2.2.2 Psychological outcomes

##### Psychological distress

Patient Health Questionnaire Anxiety and Depression Scale (PHQ-ADS; Kroenke et al., 2016). This self-report measure assesses the severity of generalised anxiety and depression symptoms. The PHQ-ADS combines the Generalized Anxiety Disorder 7-item (GAD-7; Spitzer et al., 2006) and the PHQ-9 (Kroenke Spitzer & Williams, 2001) self-report measures. The combined PHQ-ADS is a reliable and valid composite measure with good psychometric properties (Kroenke et al., 2016). Higher scores indicate higher symptoms of anxiety and depression.

##### Health anxiety

Whitley Index 7 (WI–7; Fink et al., 1999). This self-report measure includes seven items, loading on a single factor, assessing health anxiety (e.g., “Do you think there is something seriously wrong with your body?”). Participants are asked to rate to what degree each statement applies to them. The original response format (yes or no) was adapted using a 5-point Likert scale (1 = Not at all to 5 = A great deal), as recommended by Welch, Carleton and Asmundson (2009). Higher scores indicate higher levels of a health anxiety.

##### Coronavirus anxiety

Coronavirus Anxiety Scale (CAS; Lee, 2020). This self-report measure includes five items, loading on a single factor, assessing physiologically based symptoms that are aroused with COVID-19 related information and thoughts (e.g., “I felt dizzy, lightheaded, or faint, when I read or listened to news about the Coronavirus”). Participants are asked to rate how frequently they experience each anxiety symptom. The measure is scored using a 5-point time anchored scale (0 = Not at all to 4 = Nearly every day over the last 2 weeks) and scores range between 0-20. Higher scores indicate higher levels of a COVID-19 anxiety.

##### COVID-19 anxiety syndrome

COVID-19 Anxiety Syndrome Scale (C-19ASS; Nikčević & Spada, 2020). This self-report measure includes 9 items, loading on two factors, assessing features of the anxiety syndrome linked to COVID-19 including: (1) avoidance (e.g., of public transport because of the fear of contracting COVID-19); (2) checking (e.g., of symptoms of COVID-19); (3) worrying (e.g., researching symptoms of COVID-19 at the cost of other activities); and (4) threat monitoring (e.g., paying close attention to others displaying possible symptoms of COVID-19). Items relating to checking, worrying and threat monitoring load on the first factor (‘perseveration’) with the second factor consisting of avoidance items (‘avoidance’). Participants are asked to rate how frequently they experience each feature of the anxiety syndrome. The measure is scored using a 5-point time anchored scale (0 = Not at all to 4 = Nearly every day over the last 2 weeks) and scores range between 0-36. Higher scores indicate higher levels of the presence of the anxiety syndrome.

##### Work and social adjustment

Work and Social Adjustment Scale (WSAS; Mundt et al., 2002). This self-report measure assesses the severity of functional impairment and was adapted from WSAS, proposed by Mundt et al. (2002). Participants were asked to rate five items of WSAS, using a 9-point severity scale (0 = Not at all to 8 = Very severely) with the following preamble: “Thinking about the COVID-19 pandemic and the way it may have impacted your mental health please look at each statement below and rate the extent to which the following items apply to you.” Higher scores indicate higher levels of a functional impairment.

##### Personality traits

Big Five Inventory-10 (BFI-10; Rammstedt and John, 2007). This self-report measure includes 10 items, loading on five factors, assessing the Big 5 domains of personality: extraversion, agreeableness, conscientiousness, neuroticism, and openness to experience. Participants are asked to rate how well statements describe one’s personality. The measure is scored using a 5-point Likert scale (1 = Strongly disagree to 5 = Strongly agree) and scores range between 2-10 for each of the five factors. Higher scores indicate higher levels of a given personality trait.

##### Attitude towards risk and life satisfaction

These measures were adapted from a survey conducted by the German Socio-Economic Panel, which measures the risk attitudes of more than 22,000 individuals. Dohmen et al. (2011) provide evidence of the strong informational content of the questions asked in the survey by conducting an experimental verification. For example, the question about risk taking was: “How do you see yourself: are you generally a person who is fully prepared to take risks or do you try to avoid taking risks?” Participants were asked to respond, using a 10-point severity scale (0 = ‘not at all willing to take risks’ to 10 = ‘very willing to take risks’).

Using a similar scale, participants were asked to respond to the following question regarding life satisfaction: “How satisfied are you today with the following areas of your life?” The covered areas are: health, sleep, job, income, dwelling, leisure time, family life, standard of living. Higher scores indicate higher levels of satisfaction.

Finally, we also asked the following question: “Do you have the feeling that what you are doing in your life is valuable and useful?” Participants were asked to respond, using a 10-point severity scale (0 = ‘not at all valuable or useful’ to 10 = ‘completely valuable and useful’).

## 3 Empirical Results

### 3.1 Country level average scores

We start with a descriptive analysis by calculating country level average scores for the variables related to composite anxiety and depression (PHQ-ADS), health anxiety (WI-7), COVID-19 anxiety (CAS), COVID-19 anxiety syndrome (C-19ASS-P and C-19ASS-A), and work and social adjustment (WSAS). Results are reported in Table 2. The table displays a large amount of heterogeneity among Countries. We have reported in bold the entries for which a test for difference in mean scores between each Country and the rest of the sample is significant at the 1% level, after applying a Bonferroni correction to control for spurious rejections.

**Table 2:** Country level average scores with tests of difference in means.

|  | China | Germany | Italy | Sweden | UK | USA |
| --- | --- | --- | --- | --- | --- | --- |
| PHQ-ADS | **10.47** | 14.38 | **16.81** | 14.13 | **18.43** | **16.56** |
| WI-7 | **13.8** | 14.85 | 15.91 | 14.69 | **16.28** | **16.09** |
| CAS | **3.56** | 3.92 | 4.56 | **3.15** | 4.43 | **5.07** |
| C-19ASS-P | **8.17** | **8.43** | **13.4** | 9.62 | 11.14 | **12.15** |
| C-19ASS-A | **3.92** | **4.58** | **7.9** | 6.28 | **6.37** | **6.85** |
| WSAS | 22.05 | 23.63 | 23.45 | **18.86** | 22.74 | 22.59 |

*Notes:* PHQ-ADS = Patient Health Questionnaire Anxiety and Depression Scale; WI-7 = Whitley Index 7; CAS = Coronavirus Anxiety Scale; C-19ASS-P = COVID-19 Anxiety Syndrome Scale-Perseveration; C-19ASS-A = COVID-19 Anxiety Syndrome Scale-Avoidance; WSAS = Work and Social Adjustment Scale.

For PHQ-ADS, the United Kingdom, Italy, and the United States (in this order) were significantly above country level average scores, whilst China was significantly below country level average scores. With respect to health anxiety (measured by WI-7), the United Kingdom, and the United States (in this order) were significantly above country level average scores, whilst China was significantly below country level average scores.

Moving to COVID-19 specific measures, for CAS the United States alone was significantly above country level average scores whilst both Sweden and China (in this order) were significantly below country level average scores.

Disaggregating attention to COVID-19 in the two factors, namely perseverance and avoidance, gives us a similar picture. In fact, for C-19ASS-P Italy and the United States (in this order) were significantly above country level average scores whilst China and Germany (in this order) were significantly below country level average scores. With respect to C-19ASS-A Italy, the United States, and the United Kingdom (in this order) were significantly above country average scores, whilst China and Germany (in this order) were significantly below country level average scores.

Finally, for WSAS we notice a lower level of heterogeneity across Countries, with the exception of Sweden, that has recorded a markedly lower average level. This could be the consequence of the less restrictive approach adopted by the Swedish Government.

Overall, these data seem to suggest that United States, United Kingdom and Italy were consistently above average levels for both generic and COVID-19 specific measures of psychological distress, whereas at the other end of the spectrum we notice China and, to a lesser extent, Germany.

### 3.2 Regression analysis

In this Subsection we try to give an answer to two important empirical questions. First, do the differences among Countries reported in Table 2 survive when we include individual characteristics of the respondents, or the latter act as confounding factors? Secondly, what is the relationship between all the measures of psychological distress analyzed in the previous subsection, and is it possible to establish a direction of causation? We do so by specifying and estimating different regression models for each of the output variables analyzed in the previous subsection.

Because of the discreteness of data measured on a scale, we cannot rely on linear models to estimate the relationship between variables. Hence, in what follows, we resorted to ordered logit models, obtained by bucketing the different scores in categories indicating the strength of the output. The choice of an ordered logit model seems natural in this context because scores can be ordered and hence there is no arbitrariness involved.

We have grouped the output variables as follows. For the measure of composite anxiety and depression (PHQ-ADS), the thresholds came from the literature (Kroenke et al., 2016): scores below 10 were taken to indicate minimal distress, between 10 and 19 mild, between 20 and 29 moderate and finally 30 and above severe distress. For the remaining measures, as there are no predetermined available thresholds, we resorted to a data dependent approach: we chose values below the 30% quantile as a low score, between the 30% and the 70% quantile as a moderate score, and finally above the 70% quantile as a high score.^3^

Furthermore, because of the high correlation between the measures of psychological distress, we have controlled for the potential endogeneity in the ones employed as regressors, by resorting to a control function approach (for a recent survey, see Wooldridge, 2015). The approach consists in estimating the regression models in two different stages. In the first stage, we regressed the potentially endogenous variables on all the exogenous ones used in the second stage regressions, plus some regressors that we have only used in the first stage. These regressors are: personality traits, measures of satisfaction towards different aspects of life, and attitude towards risk. The *F*-statistics associated with the exogenous regressors specific to the first stage were highly significant, avoiding the problem of weak measures.^4^

In the second stage we added as extra regressors the residuals obtained from the first stage. The test of significance on the residuals can be interpreted as a test of endogeneity of the related regressor. All standard errors were obtained by 5,000 bootstrap replications. In all the regressions we included fixed effects related to religion, ethnicity, civil status, employment, education, profession, COVID-19 testing, COVID-19 vaccination status, COVID-19 death of a relative, previous mental health diagnosis, disability, critical health condition, and pregnancy. We also included control variables on weight, number of children and household size.

Results are reported in Table 3. To avoid the problem of perfect collinearity, we had to drop one of the fixed effects associated with country of residence. We have dropped the United States, so in what follows we should interpret the effect of the country of residence as being relative to the United States.

**Table 3:** Regression results.

| VARIABLES | Model (1)<br>PHQ-ADS | Model (2)<br>CAS | Model (3)<br>C-19ASS-P | Model (4)<br>C-19ASS-A | Model (5)<br>WSAS |
| --- | --- | --- | --- | --- | --- |
| PHQ-ADS |  | -0.00132 | 0.0133 | -0.0233 | 0.0716*** |
| CAS | -0.0975 |  | 0.234*** | -0.181*** | 0.242* |
| C-19ASS-P | 0.185*** | 0.446*** |  | 0.271*** | 0.171*** |
| C-19ASS-A | -0.470*** | -0.907*** | 0.627*** |  | -0.110 |
| WI-7 | 0.540*** | 0.276*** | -0.104** | 0.130*** | -0.0647 |
| Age | -0.0174*** | -0.00151 | -0.0105** | 0.00774* | 0.00659 |
| Female | 0.238*** | 0.0252 | -0.0913 | 0.0421 | -0.143** |
| Female * Age | 0.00822 | 0.00912* | -0.00881 | 0.00543 | -0.000825 |
| China | -1.261*** | -0.590 | 0.349 | -1.498*** | 0.579* |
| Germany | -0.395* | -0.191 | 0.0996 | -0.618*** | 0.803*** |
| Italy | 0.144 | 0.343* | 0.0361 | 0.239 | -0.0734 |
| Sweden | 0.114 | 0.386** | -0.488*** | 0.230 | 0.0457 |
| UK | 0.261* | -0.124 | 0.0591 | -0.0147 | 0.0578 |
| Individual Fixed Effects | YES | YES | YES | YES | YES |
| Individual Controls | YES | YES | YES | YES | YES |
| Observations | 5,049 | 5,049 | 5,049 | 5,049 | 5,049 |
| Pseudo $R^2$ | 0.390 | 0.339 | 0.341 | 0.320 | 0.191 |

Notes: PHQ-ADS = Patient Health Questionnaire Anxiety and Depression Scale; WI-7 = Whitley Index 7; CAS = Coronavirus Anxiety Scale; C-19ASS-P = COVID-19 Anxiety Syndrome Scale-Perseveration; C-19ASS-A = COVID-19 Anxiety Syndrome Scale-Avoidance; WSAS = Work and Social Adjustment Scale. \*\*\* p<0.01, \*\* p<0.05, \* p<0.1.

Starting from model (1), involving the composite measure of anxiety and depression as dependent variable (PHQ-ADS), we see that C-19ASS-P and C-19ASS-A are significant predictors, the former with a positive and the latter with a negative sign. Hence, individuals who engage with monitoring risks associated with COVID-19 are more likely to belong to groups with higher levels of PHQ-ADS. Conversely, those who avoid COVID-19 risks have a higher probability of reporting lower levels of PHQ-ADS. Predictably, WI-7 has a positive and significant effect, whereas CAS is not significant. In terms of the effect of country of residence, China confirms the findings of the previous subsection with a strongly significant negative sign, Germany (resp. UK) have marginally negative (resp. positive) signs. Moving to individual specific features, we notice that PHQ-ADS was significantly higher for younger, female, highly educated, unemployed individuals and for students.

Next, we turn our attention to the results on the three COVID-19 anxiety measures, reported in models (2) to (4). Before analysing each single measure, we notice an interesting fact: PHQ-ADS had no significant effects in any of the three models, suggesting that there was an effect of COVID-19 anxiety measures on generic measures of anxiety and depression (as per the results of model (1)), but not the other way around. This fact, consistent across all models, seems to suggest a clear direction of causation between measures of distress.

We begin with model (2), involving COVID-19 anxiety (CAS) as dependent variable, and notice a positive significant effect of WI-7 and C-19ASS-P, whilst C-19ASS-A has a negative significant effect. As for country effects, Italy confirms the findings of the previous subsection with a marginally significant sign and Sweden displays a more significant positive effect.

In model (3), where the dependent variable is C-19ASS-P, the two other COVID-19 specific measures (CAS and C-19ASS-A) display a significant positive sign; perhaps surprisingly, WI-7 has a negative sign. This finding can be reconciled with the results of model (4) below.^5^ Sweden displays a significant negative sign, probably in line with the less restrictive measures decided by the National Government to tackle COVID-19. Furthermore, C-19ASS-P was significantly lower for younger people, perhaps reflecting the less severe health risks that COVID-19 has for younger generations.

Model (4) explains the determinants of C-19ASS-A. CAS confirms the negative relationship uncovered in model (2). WI-7 and C-19ASS-P are both positive and strongly significant. This helps to explain the apparently surprising negative sign of WI-7 in model (3): individuals who are anxious about their health try to avoid so much COVID-19 related risks that they engage less in checking, worrying and monitoring. China and Germany confirm the findings of the previous subsection with negative strongly significant sign: this is in line with the efficacy with which national government measures were imposed. As for individual specific features, C-19ASS-A is significantly lower for younger people, which again could be a reflection of the lower health hazards posed by the pandemic on younger generations.

Finally, model (5) deals with WSAS, which measures the severity of functional impairment caused by COVID-19. In line with what we have observed in the previous models, individuals with high self-reported levels of CAS, C-19ASS-P and PHQ-ADS have an increased probability of belonging to groups most impaired by COVID-19. Similarly, China and Germany display a positive sign, as a reflection of the severity and reinforcement of the measures introduced by the respective national governments. Female individuals, despite displaying higher levels of psychological distress as measured by PHQ-ADS, have reported statistically significant lower levels of functional impairment.

## 4 Concluding remarks

One year into the COVID-19 pandemic, we have conducted an extensive analysis on the main determinants of four measures of psychological distress, including a composite measure of anxiety and depression (the Patient Health Questionnaire Anxiety and Depression Scale; Kroenke et al., 2016), two measures of COVID-19 anxiety (the Coronavirus Anxiety Scale; Lee, 2020 and the COVID-19 Anxiety Syndrome Scale; Nikčević & Spada, 2020; Nikčević et al., 2021; Albery, Spada, & Nikčević, 2021), and an indicator of functional impairment, using representative samples from six Countries.

Our analysis has included country and entity specific effects, hence controlling for different stages of development of the pandemic and for several individual social economic and demographic characteristics.

Our study has highlighted the following main points:

1. In the analysis of generic measures of psychological distress (such as PHQ-ASD), COVID-19 specific indeces of anxiety have proved to be an independent and important source of variation. The latter are strong determinants of PHQ-ASD, but there is no feedback effect of PHQ-ASD on either C-19ASS-P or C-19ASS-A.
2. It is useful to consider two loading factors when constructing measures of distress related to COVID-19. They capture two distinct reactions (‘perseveration’ and ‘avoidance’), and this results in opposite signs in the regression models we have estimated. Combining the two measures risks blurring these effects.
3. There is substantial heterogeneity across countries of the different measures of psychological distress surveyed. However, some of the cross-country differences highlighted in the self reported measures of psychological distress disappear when we condition on individual characteristics. This confirms the validity of a regression approach, as otherwise Country differences could arise because of omitted confounding factors.
4. Our analysis reveals that there are particular categories that have suffered the most because of COVID-19, in terms of higher scores of PHQ-ASD. Namely, younger individuals, females, students, highly educated individuals and unemployed. We believe these results provide useful evidence to national governments, in order to shape appropriate measures of support.

A natural extension to this analysis would be to determine to what extent the reported higher levels of psychological distress translate into suboptimal economic behaviour, in order to quantify a social cost of this reported phenomenon. We will investigate this in future research.

## Data Availability

The survey (upon which the database has been created) has been carried out by Qualtrics and we have signed a contract whereby we cannot share the data.

## Footnotes

1 The survey was managed by Qualtrics (www.qualtrics.com), one of the largest and most used crowd sourcing platforms for questionnaire-based studies.

2 The study was approved by Ethics Committee of Kingston University, London, United Kingdom. Participants were required to provide electronic consent and were under no obligation to complete the study. Upon completion of the questionnaires, participants were debriefed.

3 After this grouping, the dependent variable becomes the probability for a given individual to belong to a particular group, with mild, average or severe score.

4 Because of the high number of variables included in the regressions, displaying the full tables would take several pages. Therefore, we have chosen the compact format of Table 3. The complete set of results is available upon request.

5 In line with our results, Sauer et al. (2022) found no association between health worries and Covid fear in a sample of health anxious individuals. They highlighted as a possible explanation for their findings the fact that highly health anxious individuals tend to have a more restrictive self-concept only regarding themselves and not other people (Weck et al., 2012). Mental distress in COVID-19 is mostly related to distress regarding others (Mertens et al., 2020; Newby et al., 2020), and as such perseveration on COVID-related content and threat could be inversely related to the severity of health anxiety, where cognitive resources are mostly taken by worrying about the self in the context of a health condition.

